# Rare genetic variant risks in patients with sepsis-associated acute respiratory distress syndrome

**DOI:** 10.1101/2025.03.10.25323600

**Authors:** Eva Tosco-Herrera, Luís A. Rubio-Rodríguez, Adrián Muñoz-Barrera, David Jáspez, Eva Suárez-Pajes, Almudena Corrales, Aitana Alonso-González, Miryam Prieto-González, Aurelio Rodríguez-Pérez, Demetrio Carriedo, Jesús Blanco, Alfonso Ambrós, Leonardo Lorente, María M. Martín, Jordi Solé-Violán, Carlos Rodríguez-Gallego, Elena González-Higueras, Elena Espinosa, Arturo Muriel-Bombin, David Domínguez, Marina Soro, Tamara Hernández-Beeftink, José M. Añón, Jesús Villar, Beatriz Guillén-Guio, Itahisa Marcelino-Rodríguez, José M. Lorenzo-Salazar, Rafaela González-Montelongo, Carlos Flores

## Abstract

**Background:** Acute respiratory distress syndrome (ARDS) is a complex, heterogeneous, and deadly condition often resulting from pulmonary lesions due to sepsis, among other causes. There is a lack of targeted therapies to specifically treat the patients. Common genetic factors in the population (frequency >1%) have been associated with ARDS susceptibility, but systematic genetic screens of the role of rare genetic variants are lacking. We used the network of known molecular interactions to identify ARDS risks from clusters of biologically related genes containing qualifying variants (QVs) with frequency <1% likely affecting function.

**Methods:** We conducted whole-exome sequencing in sepsis patients from the GEN-SEP cohort (n=822, of which 272 developed ARDS). A network-based heterogeneity clustering algorithm was used to identify significant gene clusters (*p*<1×10^-5^). Gene-set enrichment analysis and logistic regression models aggregating QVs were used for characterization of gene clusters and findings validation.

**Results:** We identified 19 significant clusters (*p*_lowest_=3.29×10^-10^), each containing an average of 102 genes (11.6% mean similarity). QVs in eight gene clusters were associated with sepsis-associated ARDS (*p*_lowest_=2.35×10^-4^) but were not associated with 28-day survival. Clusters were enriched in several biological pathways, notably the *Interferon signaling* and *Toll-like receptor cascades*.

**Conclusions:** These results support a marked genetic heterogeneity underlying ARDS susceptibility and the presence of risk variants involving multiple biological processes that are associated with sepsis outcomes. This evidence paves the way for future development of preventive and therapeutic approaches targeting those pathways to reduce the risk for sepsis-associated ARDS.

## Introduction

Acute respiratory distress syndrome (ARDS) is a heterogeneous and complex lung condition that develops after pulmonary or systemic insults, including trauma, severe pneumonia, and sepsis (Bos & Ware, 2022). The diagnosis is based on acute onset, bilateral opacities on chest radiography or CT scan (of non-cardiac origin), and a PaO2/FiO2 ratio of less than 300 mm Hg according to the latest clinical guidelines (ARDS Definition Task Force, 2012; Grasselli et al., 2023; Matthay et al., 2024). ARDS affects almost 25% of all critically ill adult patients requiring mechanical ventilation. The overall in-hospital mortality rate exceeds 40% (Beitler et al., 2022; Bellani et al., 2016), while it increased in periods of the coronavirus disease 2019 (COVID-19) pandemic caused by severe acute respiratory syndrome coronavirus 2 (SARS-CoV-2) (Papazian et al., 2021). ARDS imposes a significant economic burden on adult intensive care unit (ICU) worldwide. However, no specific pharmacotherapy is effective for ARDS treatment (Matthay et al., 2019; Xia et al., 2023; Xu et al., 2023; Aribindi et al., 2024). Current patient management is based on life-supportive therapies for maintaining gas exchange and reduce inflammation.

ARDS develops from a hyperinflammatory state causing alveolar-capillary membrane dysregulation (Bos & Ware, 2022), leading to severe impairment of gas exchange, non-cardiogenic pulmonary edema, resulting in hypoxemia and acute respiratory failure (Zayed & Askari, 2024). Host conditions such as age, sex, comorbidities, and exposure to airborne pollutants are known risk factors for ARDS (Lemos-Filho et al., 2013; Keskinidou et al., 2022; Zhang et al., 2024). However, these factors and the clinical status poorly predict ARDS risk among patients. It is thus crucial to understand the genetic and molecular mechanisms that govern host responses to severe infections and ARDS development. This knowledge is essential for patient risk stratification and for identifying novel treatments. Host genetic factors have been increasingly recognized as affecting the severity of responses to infections. To a certain point, the genetic influence on infection severity and outcomes could be exemplified in patients with inborn errors of immunity (IEI) (Kwok et al., 2021; Casanova & Abel, 2022; Schmidt et al., 2022). Genetic studies in COVID-19 patients show that there are both common (Pairo-Castineira et al., 2023; The COVID-19 Host Genetics Initiative, 2023) and rare variants affecting gene function (Zhang et al., 2022; Matuozzo et al., 2023) associated with severe SARS-CoV-2 infections and life-threatening disease.

Genome-wide screens in cohorts of ARDS patients are still scarce in the literature (Suarez-Pajes et al., 2023). Genome-wide association studies (GWAS) in ARDS have revealed common genetic risk factors (>1% frequency) in genes involved in cellular adhesion (Christie et al., 2012), platelet activation (Bime et al., 2018), vascular permeability (Guillen-Guio et al., 2020), and immune-inflammatory processes (Du et al., 2021). Sequencing-based studies evaluating the spectrum of variant frequencies (<1%) enriched for deleterious effects have focused on small cohorts of fewer than 100 patients (Lee et al., 2012; Shortt et al., 2014; Xu et al., 2021) or on familial cases affected by IEI due to genetic defects in type I interferon signaling (Ciancanelli et al., 2015; Hernandez et al., 2018; Lim et al., 2019). The latter has also evidenced some degree of genetic overlap between IEI and ARDS (Quintanilla et al., 2021; Suarez-Pajes et al., 2023).

Given that multiple biological processes are involved in ARDS, it becomes necessary to use approaches that allow aggregating genetic information from multiple pathways to capture the genetic heterogeneity. In this study, we aimed to identify rare genetic risk factors for sepsis-associated ARDS using a gene-clustering approach. We leveraged the largest exome sequencing dataset analyzed so far for this condition and the information from biological processes for adapting to patient heterogeneity.

## Materials and methods

### Patients and ethical considerations

The study examined a cohort of patients with sepsis from different foci and etiological agents from the GEN-SEP study. These patients were enrolled between 2002 and 2019 from a network of Spanish ICUs. Except for 323 patients analyzed for the first time in this study, the rest were included in previous GWAS (Guillen-Guio et al., 2020; Hernandez-Beeftink et al., 2022). Sepsis was clinically defined according to the Third International Consensus Definitions for Sepsis (Singer et al., 2016). The clinical and demographic data are available in **Supplementary Table 1**.

Initially, 995 patients were included in the cohort, but 32 patients were excluded for having incomplete clinical data, and 141 were excluded due to the DNA quantity or integrity requirements. Consequently, 822 patients remained for whom complete whole-exome sequencing (WES) data were considered for the analyses. From those, 272 patients met ARDS criteria (ARDS Definition Task Force, 2012). These 272 patients were used as cases and 550 sepsis patients without ARDS during their hospitalization were considered as controls (**Supplementary Table 1**).

The study was reviewed by The University Hospital of the Canary Islands Review Board (CHUNSC_2021-40) and the Research Ethics Committees of participating centers, and was conducted in accordance with the Declaration of Helsinki. Written informed consents were obtained from all participants or their representatives.

### Library preparation, whole-exome sequencing, and variant calling

Genomic DNA was purified from peripheral blood samples using a commercial column-based DNA extraction kit (GE Healthcare, Chicago, IL). We measured DNA concentration using the Qubit 3.0 fluorimeter with the Qubit dsDNA High Sensitivity Assay kit (Thermo Fisher Scientific, Waltham, MA). Genomic libraries were prepared using either the DNA Prep with Enrichment kit (Illumina Inc., San Diego, CA) as described elsewhere (DíazDde Usera et al., 2020) or the SureSelect XT HS2 DNA Reagent Kit (Agilent Technologies, Santa Clara, CA). We then assessed library sizes on a 4200 TapeStation (Agilent Technologies) and determined their concentration using the Qubit dsDNA HS Assay kit (Thermo Fisher Scientific).

Sequencing of libraries was conducted at the Instituto Tecnológico y de Energías Renovables (ITER, Santa Cruz de Tenerife, Spain). Sequencing was performed in parallel along with 1% of a PhiX control V3 (Illumina Inc.) to an average depth of at least 100X on NextSeq 550, HiSeq 4000, or NovaSeq 6000 Sequencing Systems (Illumina Inc.) using 75 base pairs (bp) or 100 bp paired-end reads as recommended. We preprocessed the sequence reads using bcl2fastq v2.18 for demultiplexing, BWADMEM 0.7.15 (https://github.com/lh3/bwa) for read alignment to the GRCh37/hg19 reference, SAMtools v1.3 (http://www.htslib.org) for alignment sorting and Picard v2.10.10 (https://broadinstitute.github.io/picard/) for marking of duplicate reads as detailed elsewhere (GuillenDGuio et al., 2020). The GATK HaplotypeCaller v3.8 (https://gatk.broadinstitute.org/hc/en-us/articles/360037225632-HaplotypeCaller) was used following the Best Practices recommendations for variant calling of single-nucleotide variants (SNVs) and small insertions and deletions (indels<50 bp) with a padding of 100 bp.

Variant quality controls were applied using BCFtools v1.16 (https://github.com/samtools/bcftools) to retain only high-confidence variants in the final dataset. Variants with a PASS, depth of coverage (DP) ≥10, genotype quality (GQ) ≥20, and missingness rate (FMISS) <0.05 were included in the study. Detected variants were annotated for population allele frequency (AF) according to gnomAD Exome v2.1 (https://gnomad.broadinstitute.org), RefSeq-based annotation, functional consequences, and pathogenic potential based on ClinVar and the Gene Damage Index (GDI) <u>(Itan et al., 2015)</u>. Pathogenicity prediction scores, such as the Combined Annotation-Dependent Depletion (CADD) score v1.6, were annotated with Ensembl Variant Effect Predictor (VEP) v.105 (https://www.ensembl.org/info/docs/tools/vep/index.html) and ANNOVAR v18.04.16 (https://annovar.openbioinformatics.org/en/latest/). The in-house pipeline is fully described at https://github.com/genomicsITER/benchmarking/tree/master/WES. All analyses were carried out using the TeideHPC Supercomputing facility (http://teidehpc.iter.es/en) (see **Supplementary Methods**).

### Network-based heterogeneity clustering and biological enrichment analyses

We used the Network-based heterogeneity clustering v3 (NHC) to systematically aggregate biologically proximate genes from WES data based on a predefined protein-protein interaction network (Zhang et al., 2021). We followed recommendations provided by Zhang et al., (2021) to select qualifying variants (QVs) for analysis. These QVs had an AF ≤0.01 in non-Finnish European populations, a CADD ≥10, a GDI ≤10, and that were annotated for protein truncation prediction according to VEP (non-synonymous, missense, start/stop gain/loss, frameshift, or splicing variants). We obtained HUGO Gene Nomenclature Committee names for QVs and used them as the input for NHC on a per-sample basis. To control for genetic heterogeneity in the cohort, the first three principal components (PCs) of common variation derived from paired SNP genotyping array data (**Supplementary Methods** and **Supplementary Figure 1**) were also included in the NHC analysis.

Following recommendations (Zhang et al., 2021), gene clusters differing between cases and controls were considered significant at *p*≤1×10^-5^. A similarity matrix based on gene-level overlap was calculated among significant clusters using the Jaccard distance. Gene interaction graphs within significant clusters were represented using Cytoscape v3.10.2 (Shannon et al., 2003) to integrate the known functional interactions between gene products and the count of ARDS cases with QVs. Finally, we assessed biological pathway enrichment of each significant cluster of genes using REACTOME (https://reactome.org/). We retained enriched significant pathways (*p*≤1×10^-5^) and selected the most significant as the top pathway to be representative for each cluster (Zhang et al., 2021).

### Qualifying variant carrier associations with ARDS

We used the QVs to identify carriers among the GEN-SEP patients, provided that at least one QV was identified in any gene from a particular cluster. We restricted the analysis to significant gene clusters having minimal gene-level overlap (i.e., based on a similarity <20% and choosing the most significant cluster when compared clusters had >20%). This approach avoids an overly conservative statistical penalty due to gene-level similarity between clusters. Since only 10 of the significant clusters satisfied this condition, we applied a *p*<5×10^-3^ (*p*=0.05/10) threshold to declare that QVs carriers of a specific cluster were significantly associated with ARDS. For those 10 significant clusters, the association of QVs carriers with sepsis-associated ARDS was tested using binomial logistic regressions adjusted for sex, age, and Acute Physiology And Chronic Health Evaluation II (APACHE II) scores as in previous GWAS (Guillen-Guio et al., 2020; Hernandez-Beeftink et al., 2022) using R v4.3.2 (R Studio Team, 2020).

A schematic overview of the analysis workflow is presented in **Supplementary Figure 2.**

### Sensitivity analyses

Alternative models testing the association of QVs with ARDS risk were also conducted to assess the impact of adjusting for relevant demographic and clinical variables.

Loss-of-function variants in 15 genes of the Toll-like receptor (TLR)3- and TLR7-dependent interferon response (*IFNAR1*, *IFNAR2*, *IRF3*, *IRF7*, *IRF9*, *IKBKG*, *STAT1*, *STAT2*, *TBK1*, *TICAM1*, *TRAF3*, *UNC93B1*, *TYK2*, *TLR3*, and *TLR7*) are known to cause life-threatening COVID-19 (Matuozzo et al., 2023). We tested if ARDS risk could be explained by QVs in these genes by modelling two scenarios based on binomial logistic regressions adjusted by sex, age, and APACHE II scores: i) QVs with AF≤0.01; and ii) QVs with AF≤0.001.

To rule out that associations could be due to residual confounding by genetic heterogeneity, we devised a null model by testing the association of rare synonymous variants (AF≤0.01) with ARDS risk for the 10 significant clusters.

Finally, a Cox proportional hazards model was employed with a subset of 665 patients for which follow-up survival data was available (152 deaths and 513 survivors) to test associations between QVs carriers at those 10 gene clusters and 28-day survival using the *survival* v3.7-0 R package. We did this to rule out the possibility that the QVs carrier status was associated with mortality rather than ARDS. Two alternative survival models were used for this: i) model 1 adjusted for sex, age, and APACHE II; and ii) model 2 adjusted for sex, age, APACHE II, and ARDS status of the patient. The proportionality of Hazard Ratios (HR) assumption across covariates was verified using scaled Schoenfeld residuals. For gene clusters that did not fulfill this assumption, the *coxphw* v4.0.3 R package was applied to perform a weighted estimation of average HR and avoid the need for HR proportionality across time.

## Results

### NHC analysis of exomes in ARDS patients compared to sepsis controls

The NHC analysis revealed 19 gene clusters significantly different between ARDS cases and sepsis controls (*p*-value range= 3.29×10^-10^ to 5.68×10^-6^) (**Table 1**, **Figure 1**). Significant clusters contained an average of 102 genes and had a mean gene-level similarity of 11.6% with other clusters (standard deviation [SD]= 22.6%) (**Supplementary Figure 3**). None of the genes identified in the previous sequencing-based studies of ARDS cohorts (Lee et al., 2012; Shortt et al., 2014; Xu et al., 2021) were present in the significant gene clusters.

**Figure 1.**
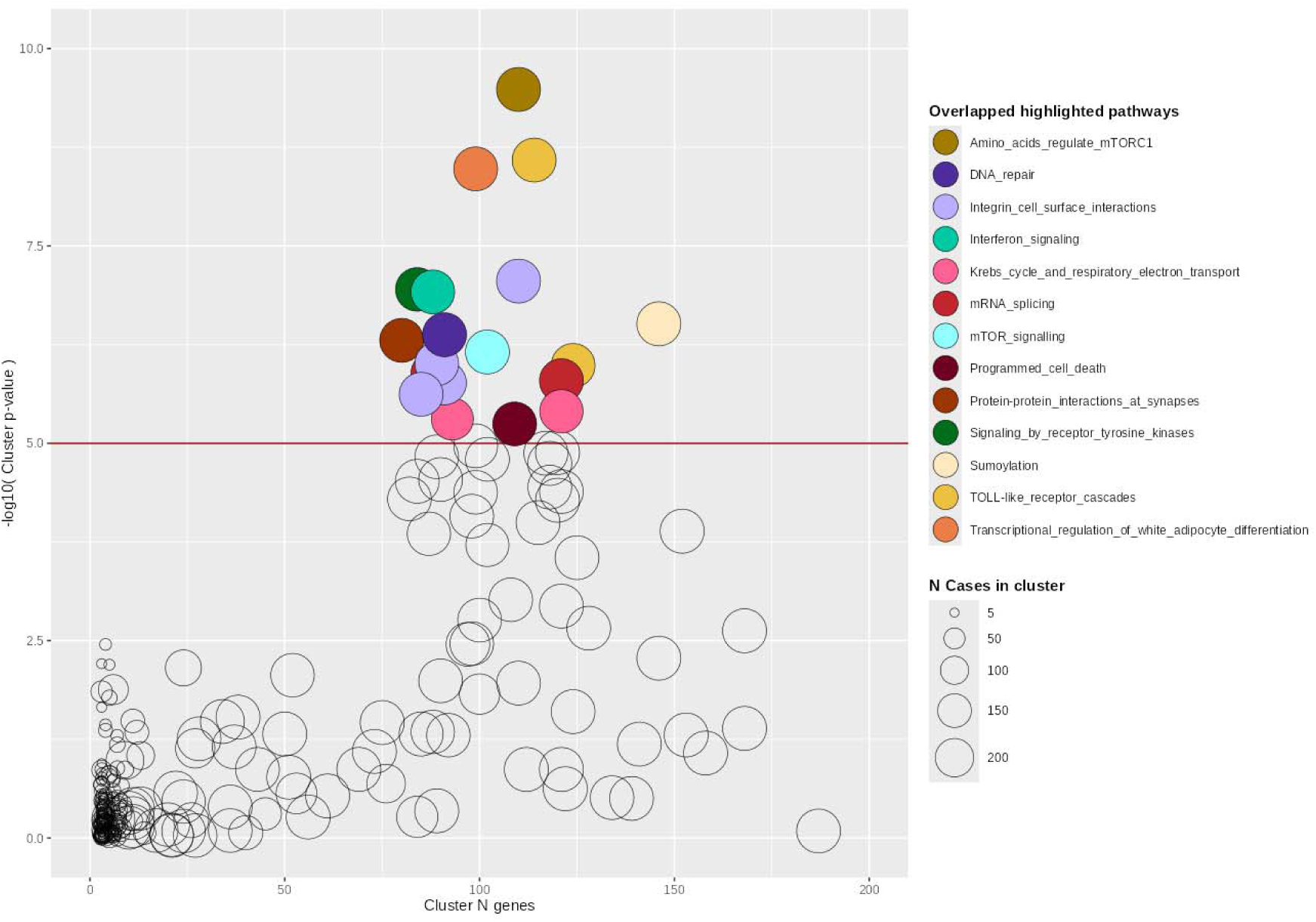
Bubble plot of gene clusters identified by the Network-based Heterogeneity Clustering analysis of ARDS cases compared to sepsis controls. Gene clusters are represented by bubbles, where the size of each is proportional to the number of ARDS cases assigned. Colors denote significant clusters (p≤1×10^-5^) that share the top significant pathway. The x-axis represents the number of genes included in each cluster, and the y-axis represents the significance (-log_10_[*p*-value]).

**Table 1.**
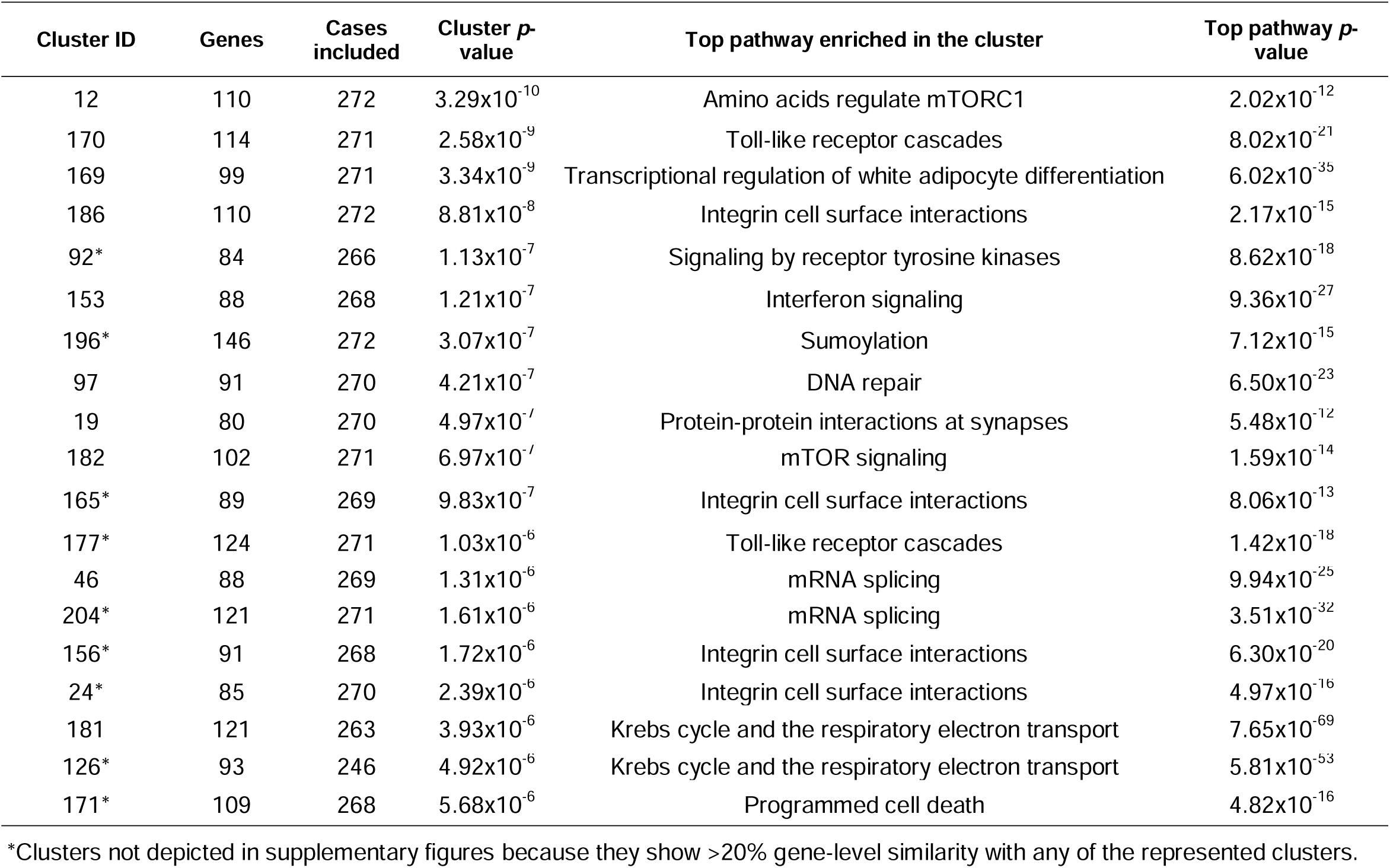
Gene clusters significantly differing between ARDS cases and sepsis controls.

The most frequent pathways among the clusters (**Supplementary Table 2**) were *Integrin cell surface interactions* (clusters 24, 156, 165, and 186) (**Supplementary Figure 4**), *TLR cascades* (clusters 170 and 177) (**Supplementary Figure 5)**, and the *Krebs cycle and the respiratory electron transport* (clusters 126 and 181) (**Supplementary Figure 6**). Less frequently enriched pathways were *Transcriptional regulation of white adipocyte differentiation* (cluster 169) (**Supplementary Figure 7**), *Interferon signaling* (cluster 153) (**Supplementary Figure 8**), and *mTOR signaling* (cluster 182) (**Supplementary Figure 9**). Additional information can be found in **Supplementary Figures 10-13**.

We identified 208 genes (or “nodes” in the networks) exhibiting QVs in ≥10 cases (average of 15.6 [SD=6.79] cases) in the 19 gene clusters (**Supplementary Table 3**). Of those, 25 (12.02%) are considered causes of IEIs according to the 2024 classification update (https://iuis.org/committees/iei/). The top 10 genes accumulating QV carriers among ARDS cases were: *CFTR* (57 cases), *MPDZ* (56 cases), *SRRM2* (38 cases), *BRCA2* (33 cases), *NOTCH2* (32 cases), *PLCG2* (32 cases), *CGN* (32 cases), *PLXNA4* (32 cases), *NRP2* (31 cases), and *TJP2* (31 cases). Most of these genes were part of more than one significant cluster (*CFTR* in 13/19, *NOTCH2* and *CGN* in 4/19, *SRRM2, PLXNA4, BRCA2* and *NRP2* in 2/19). *MPDZ*, *PLCG2*, and *TJP2* were each part of one significant cluster (**Supplementary Table 4**).

### Qualifying variant carriers and sepsis-associated ARDS risk

We then tested the association between carrier status for the QVs in the 10 gene clusters having the lowest pairwise similarity with ARDS risk using regression models. Carrier status in eight of the gene clusters was associated with increased risk of sepsis-associated ARDS (**Table 2**), reflecting the broad systemic response involved in ARDS development. Two gene clusters failed to provide an estimate of the effect size due to model convergence problems. These results were robust to model adjustments for relevant demographic and clinical variables (**Supplementary Table 5**). Null models with the synonymous variants for the 10 gene clusters at the same AF threshold showed no association with ARDS (**Supplementary Table 6**).

**Table 2.** Association of qualifying variant carriers with sepsis-associated ARDS risk in the 10 significant gene clusters with the lowest pairwise similarities.

| Cluster ID | Top enriched pathway | Genes | Odds Ratio (95% CI) | p-value |
| --- | --- | --- | --- | --- |
| 12 | Amino acids regulate mTORC1 | 110 | NAN<sup>†</sup> | - |
| 170 | Toll-like receptor cascades | 114 | 30.1 (4.1-222) | 8.17x10<sup>-4</sup> |
| 169 | Regulation of white adipocyte differentiation | 99 | 25.3 (3.46-185) | 1.45x10<sup>-3</sup> |
| 186 | Integrin cell surface interactions | 110 | NAN<sup>†</sup> | - |
| 153 | Interferon signaling | 88 | 7.1 (2.53-19.9) | 1.96x10<sup>-4</sup> |
| 97 | DNA repair | 91 | 12.1 (2.89-50.4) | 6.37x10<sup>-4</sup> |
| 19 | Protein-protein interactions at synapses | 80 | 26.2 (3.54-194) | 1.38x10<sup>-3</sup> |
| 182 | mTOR signaling | 102 | 11.1 (2.63-46.6) | 1.05x10<sup>-3</sup> |
| 46 | mRNA splicing | 88 | 8.66 (2.65-28.3) | 3.50x10<sup>-4</sup> |
| 181 | Krebs cycle and respiratory electron transport | 121 | 3.91 (1.89-8.08) | 2.35x10<sup>-4</sup> |

CI: confidence interval. <sup>†</sup> Models not converging, attributable to disproportionate distribution of carriers and non-carriers and leading to unreliable effect size estimates.

The most significant association was found for carriers of QVs in genes from cluster 153, where *Interferon signaling* was the top-most enriched pathway (Odds Ratio [OR]=7.1; 95% Confidence Interval [CI]=2.53-19.9; *p*-value=1.96×10^-4^). However, the largest effect size was found for carriers of QVs in genes from cluster 170, in which *TLR cascades* was the top enriched pathway (OR=30.1; 95%CI=4.1-222; *p*-value=8.17×10^-4^). Associations were not significant for models that only considered QV carriers in the TLR3- and TLR7-dependent interferon response genes underlying life-threatening COVID-19 at AF≤0.01 (OR=0.82; 95%CI=0.56-1.19; *p*-value=0.288) or at AF≤0.001 (OR=0.79; 95%CI=0.49-1.26; *p*-value=0.317).

Finally, to rule out that QVs carrier status in these gene clusters was merely reflecting higher mortality, associations between QVs carriers and 28-day survival were tested using two alternative models. Models for all but two of the gene clusters (clusters 46 and 181) passed the proportionality of hazards test and none of the models were significant (**Table 3**). This evidenced that associations of carrier status with ARDS risk were not confounded by mortality.

**Table 3.** Results of the 28-day survival analyses for the 10 significant gene clusters with the lowest pairwise similarities.

| Cluster ID | Top enriched pathway | Genes | Model 1: sex, age, APACHE II | Model 2: sex, age, APACHE II, ARDS status |
| --- | --- | --- | --- | --- |
|  |  |  | Hazard Ratio (95% CI), *p*-value | Hazard Ratio (95% CI), *p*-value |
| 12 | Amino acids regulate mTORC1 | 110 | 1.24 (0.61-2.53), *p*=0.552 | 1.04 (0.50-2.17), *p*=0.909 |
| 170 | Toll-like receptor cascades | 114 | 3.20 (1.18-8.64), *p*=0.022 | 2.79 (1.02-7.62), *p*=0.045 |
| 169 | Regulation of white adipocyte differentiation | 99 | 1.01 (0.51-1.98), *p*=0.986 | 0.86 (0.43-1.72), *p*=0.671 |
| 186 | Integrin cell surface interactions | 110 | 1.08 (0.48-2.46), *p*=0.845 | 0.92 (0.40-2.11), *p*=0.842 |
| 153 | Interferon signaling | 88 | 0.80 (0.45-1.43), *p*=0.455 | 0.70 (0.39-1.27), *p*=0.243 |
| 97 | DNA repair | 91 | 1.07 (0.52-2.20), *p*=0.847 | 0.92 (0.44-1.91), *p*=0.824 |
| 19 | Protein-protein interactions at synapses | 80 | 4.80 (1.19-19.37), *p*=0.028 | 4.14 (1.02-16.86), *p*=0.047 |
| 182 | mTOR signaling | 102 | 1.42 (0.58-3.46), *p*=0.444 | 1.21 (0.49-2.99), *p*=0.679 |
| 46<sup>#</sup> | mRNA splicing | 88 | 1.17 (0.54-2.56), *p*=0.684 | 1.05 (0.48-2.31), *p*=0.896 |
| 181<sup>#</sup> | Krebs cycle and respiratory electron transport | 121 | 1.53 (0.80-2.92), *p*=0.197 | 1.39 (0.73-2.67), *p*=0.318 |

APACHE II: Acute Physiology and Chronic Health Disease Classification System II. CI: confidence interval. Analysis performed in 665 of the patients with available 28-day mortality data. <sup>#</sup>Used the weighted Cox regression model, not assuming the proportionality of hazards.

## Discussion

Studies performed over the last 30 years have revealed that a fraction of severe infectious disease by a particular microorganism can be etiologically explained by human genetic and immunological determinants in a growing proportion of patients (Casanova & Abel, 2024). Although most of this evidence is based on familial analyses or isolated cases, cohort studies also support this idea, and the COVID-19 pandemic has accelerated the acceptance of this view by a wider audience (Casanova & Abel, 2022). This is consistent with the idea that there is a strong genetic influence on the risk of fatal outcomes from infections, as shown by family, twin, and adoption studies (Petersen et al., 2010; Miller et al., 2021). Here, we show that genetic variation affecting biologically proximal gene clusters is associated ARDS risk among sepsis patients. This provides novel, actionable evidence for specific therapies targeting host dysregulation. We identified QVs with predicted impact in gene function in a cohort of unrelated sepsis patients. We used a network-based approach to identify variant enrichment in high-confidence, biologically connected gene clusters. The most salient genetic associations, by effect size estimates or by the statistical significance, were observed for variants in gene clusters enriched in the *TLR cascades* and the *Interferon signaling*, respectively. We also show that there is no overlap between the genetic risk for ARDS and patient survival, likely indicating that sepsis severity and mortality may be governed by distinct genetic factors, as proposed for COVID-19 (Guillen-Guio et al., 2024; Minnai et al., 2024). To our knowledge, this is the first study reporting that rare genetic variation is associated with sepsis-associated ARDS.

Analyses accounting for aggregated variants of genes and pathways, as opposed to traditional GWAS frameworks with variant- or gene-level analyses, have shown clear power improvements. These enable capturing the genetic architecture of complex traits, particularly affected by genetic heterogeneity and incomplete penetrance, and for providing biological explanations of disease risk and novel therapeutic targets (Zhang et al., 2021; Smith et al., 2022). Similar concepts underlie genetic screenings in Mendelian diseases when prioritizing the genetic etiology from WES data based on protein interaction networks combined with phenotype information (Smedley et al., 2015). Our study demonstrates that these approaches are powerful for assessing the heterogeneous genetic risks of sepsis-associated ARDS. We have identified that aggregated gene variant effects of *TLR cascades* and *Interferon signaling* are strongly associated. While the gene cluster enriched in *Interferon signaling* was associated with smaller effect size, previous studies have found significant discrepancies of effect size estimates derived from models using predicted *vs.* biochemically ascertained loss-of-function variants (Boos et al., 2024). Thus, the aggregated estimated effect sizes reported in this study should be interpreted with caution and require validation in independent studies.

Common genetic variation in stress and immune response-related genes, particularly those involved in pathogen recognition and signaling (Casanova & Abel, 2022), influence individual responses to infection and organ dysfunction (Burnham et al., 2024). TLR-mediated activation is intrinsically related to immune response to pathogens. Experimental animal models of sepsis support that *TLR cascades* are determinants of survival (Beutler et al., 2003). Myd88 or Tlr4 knock-outs, as well as neutralization of TLR-4, protects mice from lethal Gram-negative sepsis (Roger et al., 2009), suggesting that excessive inflammation mediated by *TLR cascades* contributes to sepsis aggravation. This is in line with the prevailing view that sepsis and sepsis-associated organ dysfunction are due to a dysregulated immune response dependent on different factors, including innate immune activation (Meyer & Prescott, 2024). Variants from several of the genes participating in the *TLR cascades* have been associated with differential cytokine responses to pathogen associated molecular patterns in humans (Wurfel et al., 2008). Patients with inherited deficiencies in MyD88 and IRAK-4, which are crucial for canonical TLR signaling, are particularly prone to specific invasive bacterial infections. Remarkably, clinical and laboratory signs of inflammation develop slowly in these patients, even during invasive bacterial disease (Picard et al., 2010). Common variants in *TLR1* have been linked to increased risk of ARDS, organ dysfunction, and TLR1-mediated production of inflammatory cytokines in healthy volunteers. Those variants have also been associated with sustained pro-inflammatory responses in sepsis patients (Wurfel et al., 2008; Pino-Yanes et al., 2010). One of the main etiological factors revealed so far of life-threatening viral infections is the type-I interferon response genes (Casanova & Abel, 2020; Matuozzo et al., 2023). There is evidence that common and rare loss-of-function variants of genes involved in TLR-3, TLR-7, and IRF7-dependent type I interferon immunity are key components associated with severe COVID-19 (Zhang et al., 2020; Pairo-Castineira et al., 2023), and patients with TLR3 deficiency are particularly susceptible to herpes simplex encephalitis and prone to critical influenza pneumonia (Casanova & Abel, 2024). In COVID-19, rare variant aggregated effects have shown very strong (OR >20) effect sizes in independent cohorts (Matuozzo et al., 2023; Boos et al., 2024). Despite models focusing on TLR3- and TLR7-dependent interferon response genes underlying life-threatening COVID-19 did not explain ARDS risk in our study, our results support a highly heterogeneous genetic component associated with rare variants affecting genes of the *TLR cascades* and *Interferon signaling* and with strong aggregated effect sizes in ARDS among non-viral sepsis patients given that most cases with pathogens identified in the cohort (>90%) were due to bacterial infections.

Our results also identified associations between other gene clusters and pathways that seem to play a key role in ARDS risk. However, their pathogenic links are less frequently explored in the literature and are further blurred by the pervasive genetic pleiotropy underlying complex diseases (Gratten & Visscher, 2016), the important genetic heterogeneity of ARDS, and the profound systemic nature of the underlying condition (Grasselli et al., 2023). The gene cluster enriched in *Integrin cell surface interactions* is significant at pathophysiological level in the detrimental fibroproliferative response occurring in ARDS (Burnham et al., 2014). This view is supported by the shared genetic component, particularly of genes in integrin and cadherin signaling pathways, between idiopathic pulmonary fibrosis and severe COVID-19 (Guillen-Guio et al., 2024). Carriers of rare variants affecting *CFTR* gene function, who do not develop cystic fibrosis, have a higher risk of severe COVID-19 symptoms and death (Baldassarri et al., 2021). *CFTR* heterozygous carriers are at higher risk of respiratory infections (Polgreen et al., 2018; Miller et al., 2020), and a common intronic risk variant in *CFTR* is associated with respiratory failure due to infections (Chen et al., 2021). *CFTR* was found in 13 out of the 19 significant gene clusters revealed in our study. Interestingly, it has been recognized as a possible therapeutic target in ARDS due to severe pneumonia (Frick, 2023) based on findings in experimental animal models (Honrubia et al., 2023). Increasing evidence shows that cell-cell signaling mechanisms play an important role during inflammation (Aman & Margadant, 2023). On the other hand, the acute and energetic early host defense associated with ARDS is known to lead to metabolic failure, immune cell dysfunction (Cheng et al., 2016), and immunosuppression (Meyer & Prescott, 2024), where mitochondrial dysfunction is central (Supinski et al., 2020). The reasons are diverse and related to the key role of mitochondria in supporting energy-dependent processes through cellular respiration, linked to the *Krebs cycle and respiratory electron transport*, and their metabolic influence on immune response (Nedel et al., 2024). These and the other significant gene clusters revealed, which are enriched in intertwined biological processes, suggest the possibility that multiple and biologically related genetic factors are acting in combination contributing to ARDS risk.

We acknowledge several limitations in the study. First, we support the biological relevance of our findings with the published literature, but we lack data from similar sequencing studies in independent sepsis-associated ARDS patients that can validate our findings. Most published sequencing studies to date have focused on patients with COVID-19 or on kindreds with affected family members. Second, since the GEN-SEP study only represents individuals of European genetic ancestry, it is difficult to predict how our findings generalize to other populations. Third, given the large heterogeneity of ARDS, we relied on a network-based approximation to identify multiple genes that are biologically connected and that could be collectively associated with the disease risk. Thus, while our analyses were unrestricted to candidate genes or inheritance models and offers an unbiased genome-wide assessment, we cannot draw conclusions at variant- or gene-level data. Lastly, our analyses relied on QVs that were kept in the study based on their frequency and potential protein truncation prediction. We do not provide biochemical evidence of QVs effects in gene function, which could have improved the prioritization of perturbed gene clusters and refined the most salient pathways associated with ARDS.

## Conclusions

Our findings highlight the complexity of the genetic factors involved in sepsis-associated ARDS risk. These factors affect diverse biological processes, including *Interferon signaling* and *TLR cascades*, that may reconcile with its complex pathophysiology. They also support an underlying landscape of genetic heterogeneity in the spectrum of rare variants influencing disease susceptibility. It is likely that some of these genetic factors could be actionable, offering a possibility to identify therapeutic targets for further research to mitigate the risk of severe outcomes among sepsis patients.

## Acknowledgements

We would like to acknowledge Dr. Peng Zhang for his availability and assistance with cluster network visualization and variant-to-gene mapping, integrated into the NHC v3 update. The authors would like to thank the TeideHPC Supercomputer facility team (https://teidehpc.iter.es/en/home) for their support, as well as to the members from the Genomics Division at the Instituto Tecnológico y de Energías Renovables (ITER) for their continuous support. E.T.-H, L.A.R.-R. and E.S.-P. acknowledge the training support provided by the University of La Laguna.

## Author Contributions

Conceptualization, C.F.; Data curation, E.T.-H., L.R.-R., D.J., E.S.-P., A.C., T.H.-B., B.G.-G, R.G-M., J.M.L.-S.; Visualization, E.T.-H., D.J.; Writing—original draft preparation, E.T.-H., I.M.-R., C.F.; Writing—review and editing, all authors; Supervision, C.F.; Funding acquisition, C.F. All authors have read and agreed to the published version of the manuscript.

## Data Availability Statement

Raw genotype or phenotype data cannot be made publicly available due to restrictions imposed by the ethics approval.

