## Supplementary material for "Rare genetic variant risks in patients with sepsis-associated acute respiratory distress syndrome"

Supplementary Methods	2

Summary of the whole-exome sequencing data of the GEN-SEP cohort	2

SNP array data from the GEN-SEP cohort	2

Supplementary references	2

Supplementary Tables	4

Supplementary Table 1.	4

Supplementary Table 6.	5

SUPPLEMENTARY FIGURES	6

Supplementary Figure 1.	6

Supplementary Figure 2.	7

Supplementary Figure 3.	8

Supplementary Figure 4.	9

Supplementary Figure 5.	10

Supplementary Figure 6.	11

Supplementary Figure 7.	12

Supplementary Figure 8.	13

Supplementary Figure 9.	14

Supplementary Figure 10.	15

Supplementary Figure 11.	16

Supplementary Figure 12.	17

Supplementary Figure 13.	18

### Supplementary Methods

#### **Summary of the whole-exome sequencing data of the GEN-SEP cohort**

Patient samples were sequenced to a mean target coverage of 105X. An average of 123 million reads per patient were captured, where 85% of the on-target regions were covered at 30X depth, and an average of 122.7 million reads (99.6%) were aligned to the reference. In total, an average of 73,705 SNVs and 13,138 small indels per sample were called with an average variant depth of 70.3X. Based on these, the estimated average transition-to-transversion ratio was 2.35, close to the expected range for whole-exome data (Trubetskoy et al., 2015), considering that the addition of a 100 bp padding extends the captured regions into intronic sequences, thereby increasing both the total sequence coverage and total number of called variants.

#### **SNP array data from the GEN-SEP cohort**

SNP genotyping of samples was completed with the Axiom® Genome-Wide Human CEU 1 Array (Thermo Fisher Scientific, Waltham, MA) at the National Genotyping Centre (CeGen) as described elsewhere (Guillen-Guio et al., 2020; Hernandez-Beeftink et al., 2022). Quality controls were performed using PLINK v1.90 and R v4.3.2 (R Studio Team, 2020). Samples with missing clinical information, genotype call rates (CR) <95%, sex mismatches between genetic inferred sex and the clinical data, samples with high degree of kinship (PIHAT>0.2), and heterozygosity outliers were excluded. Variants with minor allele frequency <1%, genotype completion rate <95%, or strongly deviating from the Hardy Weinberg equilibrium expectations (*p*<1.0x10^-6^) were excluded from the analyses. For principal components (PCs) analysis, we removed the variants located in regions known to exhibit long-range linkage disequilibrium (LD) and also pruned the variants in high LD using the function “indep-pairwise” of PLINK v1.9, setting a r^2^ of 0.15 to keep approximately 100,000 independent variants. The first three PCs were derived with PLINK and used for downstream analyses.

### Supplementary Tables

| **Supplementary Table 1.** Demographics and clinical characteristics of the patients from the GEN-SEP cohort included in this study. | | | | |
| --- | --- | --- | --- | --- |
| Variable | All (n=822) | ARDS cases (n=272) | Sepsis controls (n=550) | p-value^#^ |
| Sex, males (%) | 523 (63.63) | 187 (68.75) | 336 (61.09) | 0.0317 |
| Age, mean (SD) | 63.18 (15.05) | 62.17 (13.91) | 63.68 (15.58) | 0.0743 |
| BMI, mean (SD) | 27.43 (6.06) | 28.65 (6.77) | 26.79 (5.55) | 5.69x10^-3^ |
| SOFA, median (IQR) | 8 (6-11) | 9 (7-12) | 8 (6-10) | 4.52x10^-7^ |
| APACHE II, median (IQR) | 19 (15-24) | 21 (16-27) | 18 (14-23) | 9.88x10^-7^ |
| Days from diagnosis, median (IQR) | 20 (11-37) | 24 (13-49.5) | 18.5 (10-31.8) | 2.52x10^-3^ |
| ICU mortality, count (%) | 199 (24.21) | 99 (36.4) | 100 (18.18) | 1.76x10^-9^ |
| 28-day mortality, count (%) | 155 (18.86) | 71 (26.10) | 84 (15.27) | 7.79x10^-5^ |
| Source of infection, count (%) |  |  |  | 5.32x10^-9^ |
| Extrapulmonary | 486 (59.12) | 126 (46.32) | 360 (65.45) |  |
| Pulmonary | 237 (28.83) | 113 (41.54) | 124 (22.55) |  |
| Pathogen identified, count (%) |  |  |  | 0.0432 |
| Gram(+) | 146 (17.76) | 51 (18.75) | 95 (17.27) |  |
| Gram(-) | 198 (24.09) | 63 (23.16) | 135 (24.54) |  |
| Gram(+) and Gram(-) | 78 (9.49) | 19 (6.99) | 59 (10.73) |  |
| Fungi | 13 (1.58) | 3 (1.10) | 10 (1.82) |  |
| Viral | 19 (2.31) | 12 (4.41) | 7 (1.27) |  |
| Polymicrobial | 61 (7.42) | 19 (6.98) | 42 (7.64) |  |
| Unidentified | 307 (37.35) | 105 (38.60) | 202 (36.73) |  |
| Patients with comorbidities*, count (%) | 382 (46.47) | 120 (44.12) | 262 (47.64) | 0.258 |
| APACHE-II: Acute Physiology and Chronic Health Disease Classification System II; BMI: Body mass index; ICU: Intensive care unit; IQR: Interquartile range; SD: Standard deviation; SOFA: Sequential Organ Failure Assessment Score; ^#^Used a chi-squared test for comparisons, except for age, BMI, SOFA, APACHE II, and days from diagnosis for which we used the Mann-Whitney U-test. *Comorbidities include: age ≥ 80 years, pregnancy, alcoholism and smoking habits, or personal history of respiratory, cardiovascular, metabolic, endocrine, oncological, hematological, autoimmune, connective tissue, renal, hepatic, neurological or infectious conditions. | | | | |

| **Supplementary Table 6.** Results of the null model based on logistic regression analysis of rare (AF≤0.01) synonymous variants in gene clusters with minimal similarity. The models were adjusted for sex, age and APACHE II scores. | | |
| --- | --- | --- |
| **Cluster ID** | **Top enriched pathway** | **Odds Ratio (95% CI) p-value** |
| 12 | Amino acids regulate mTORC1 | 1.27 (0.79-2.04) p=0.320 |
| 170 | Toll-like receptor cascades | 1.56 (0.83-2.95) p=0.170 |
| 169 | Regulation of white adipocyte differentiation | 1.66 (0.87-3.17) p=0.120 |
| 186 | Integrin cell surface interactions | 1.03 (0.57-1.85) p=0.920 |
| 153 | Interferon signaling | 0.94 (0.55-1.60) p=0.810 |
| 97 | DNA repair | 0.84 (0.48-1.49) p=0.550 |
| 19 | Protein-protein interactions at synapses | 0.90 (0.61-1.33) p=0.590 |
| 182 | mTOR signaling | 2.16 (0.71-6.55) p=0.170 |
| 46 | mRNA splicing | 0.66 (0.435-0.99) p=0.048 |
| 181 | Krebs cycle and respiratory electron transport | 0.80 (0.55-1.16) p=0.230 |
| **Supplementary Figures**  **Supplementary Figure 1.** Plot of the first two principal components (PCs) of genetic variation of GEN-SEP patients and of data from the 1000 Genomes Project from African (AFR), American (AMR), East Asian (EAS), European (EUR), and South Asian (SAS) populations. The percentages represent the explained variability of each axis.  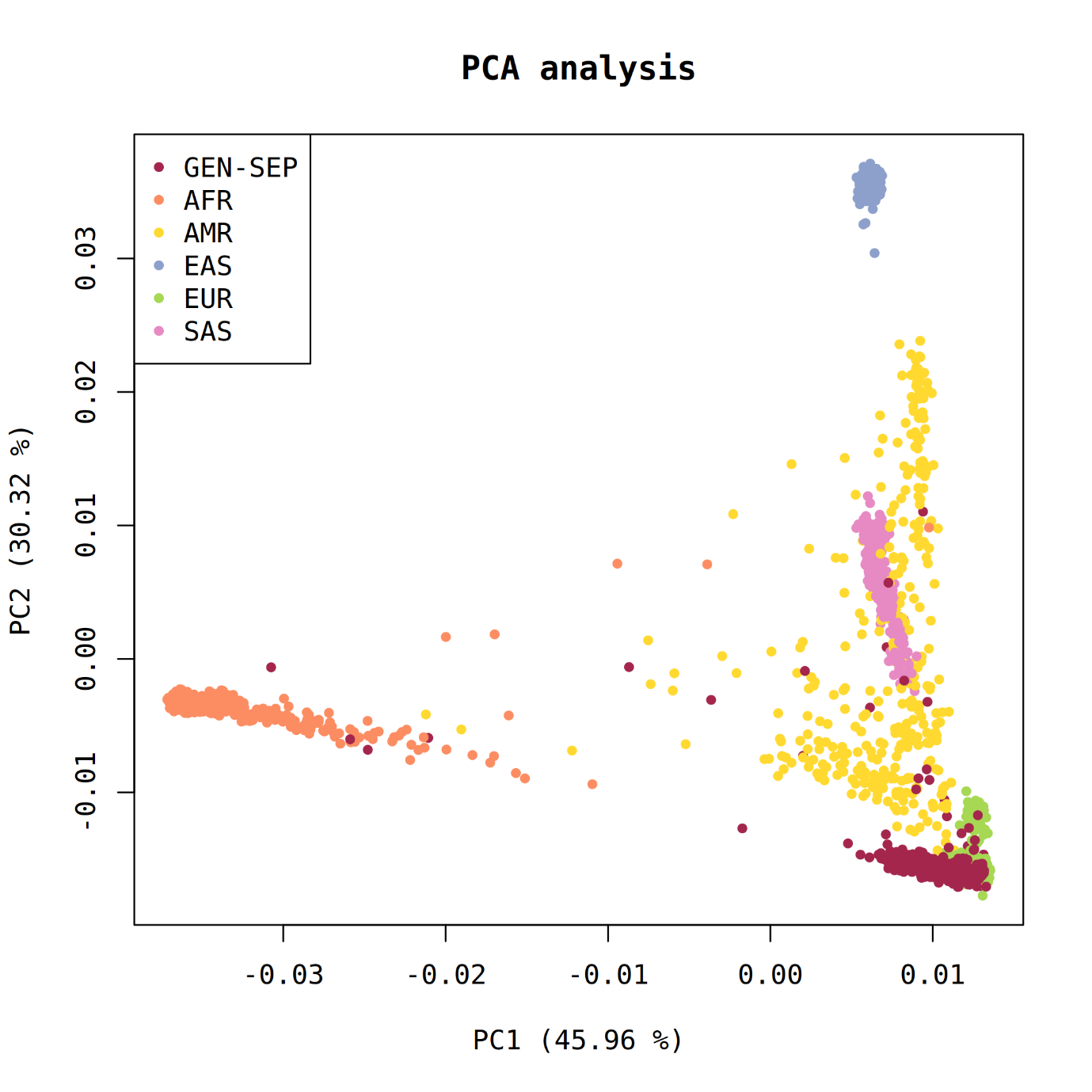 | | |

#### **Supplementary Figure 2.** Schematic overview of the sequencing workflow, filtering for qualifying variant identification, and the network-based heterogeneity clustering (NHC) analysis.

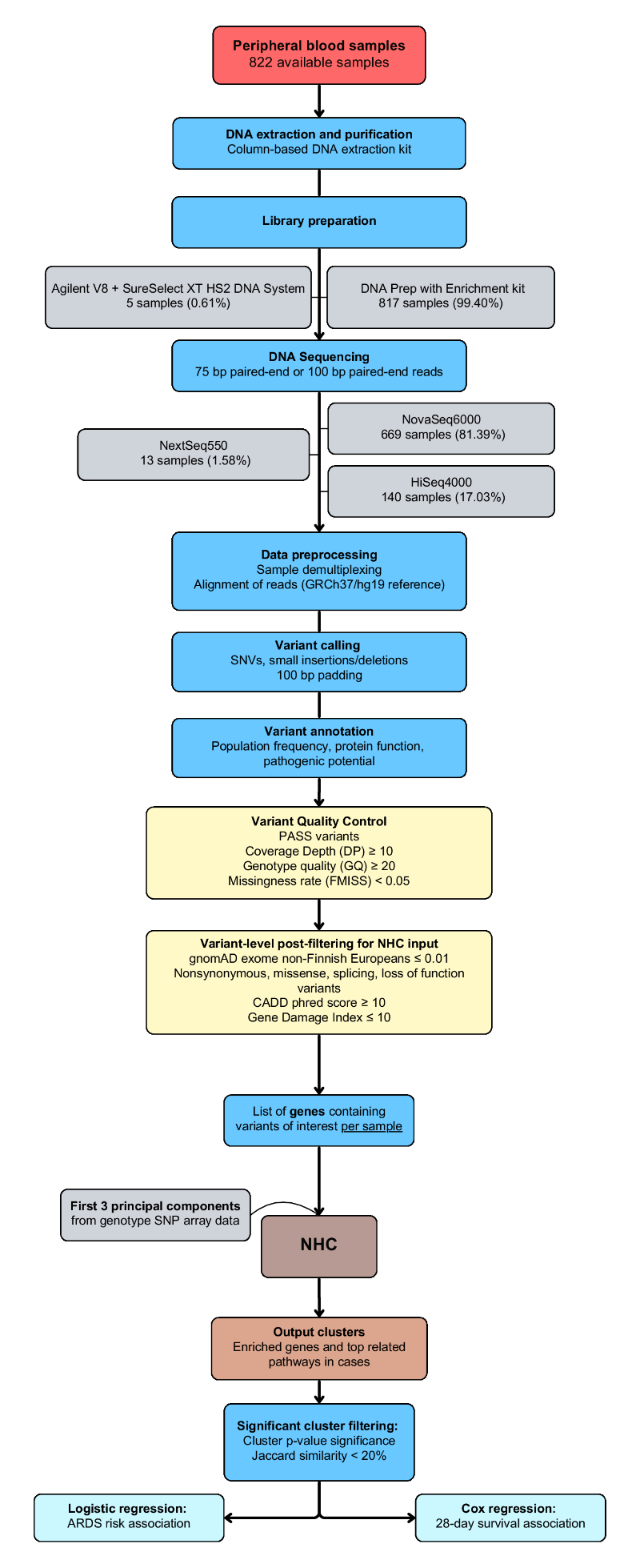

#### **Supplementary Figure 3.** Similarity matrix, as the inverse of the Jaccard distance, reflecting the percentage of gene-level overlap between significant gene clusters. Similarity between clusters is expressed in percentage (%) and by the color intensity of each box. Clusters with similarity >20% are highlighted in bold.

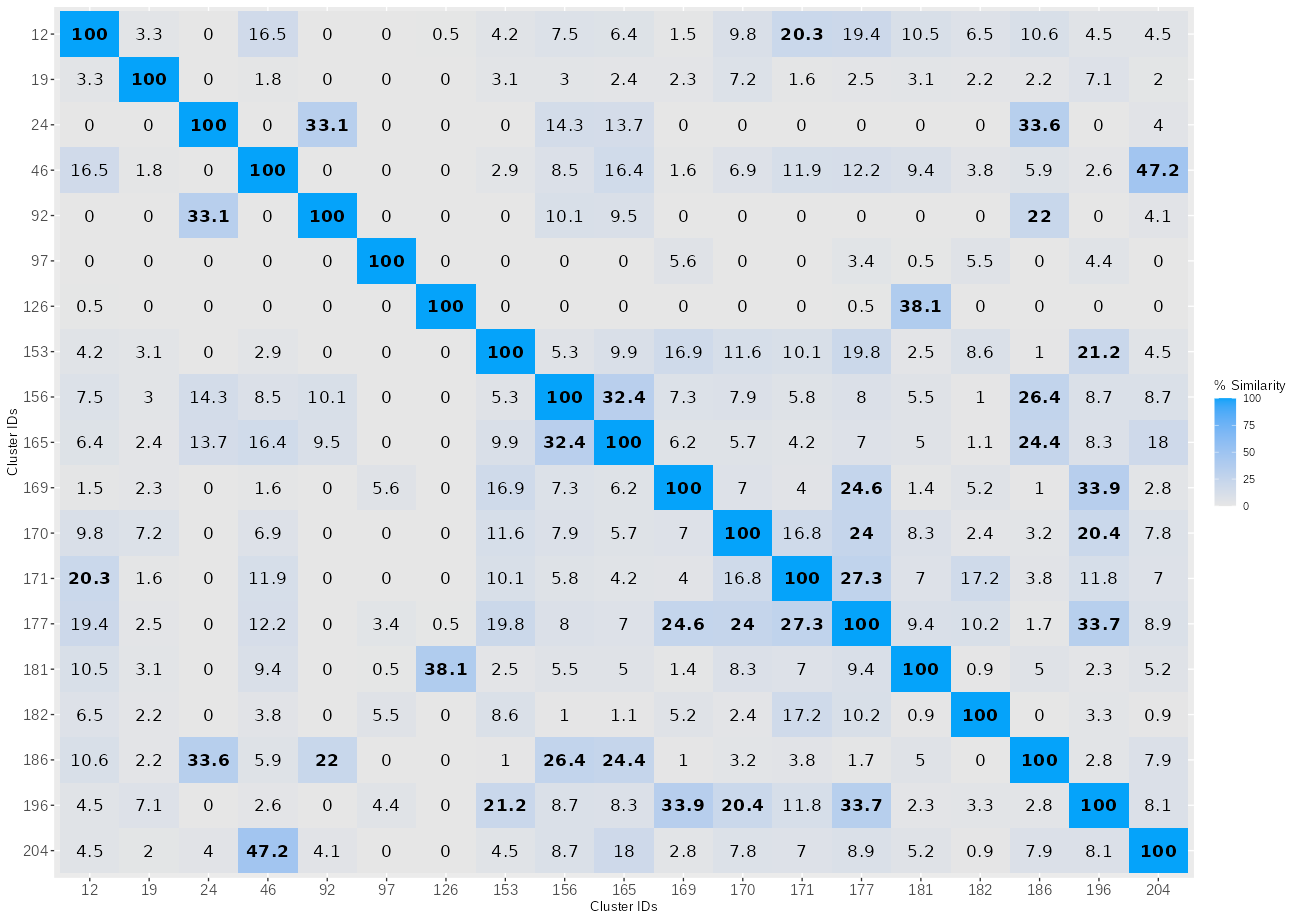

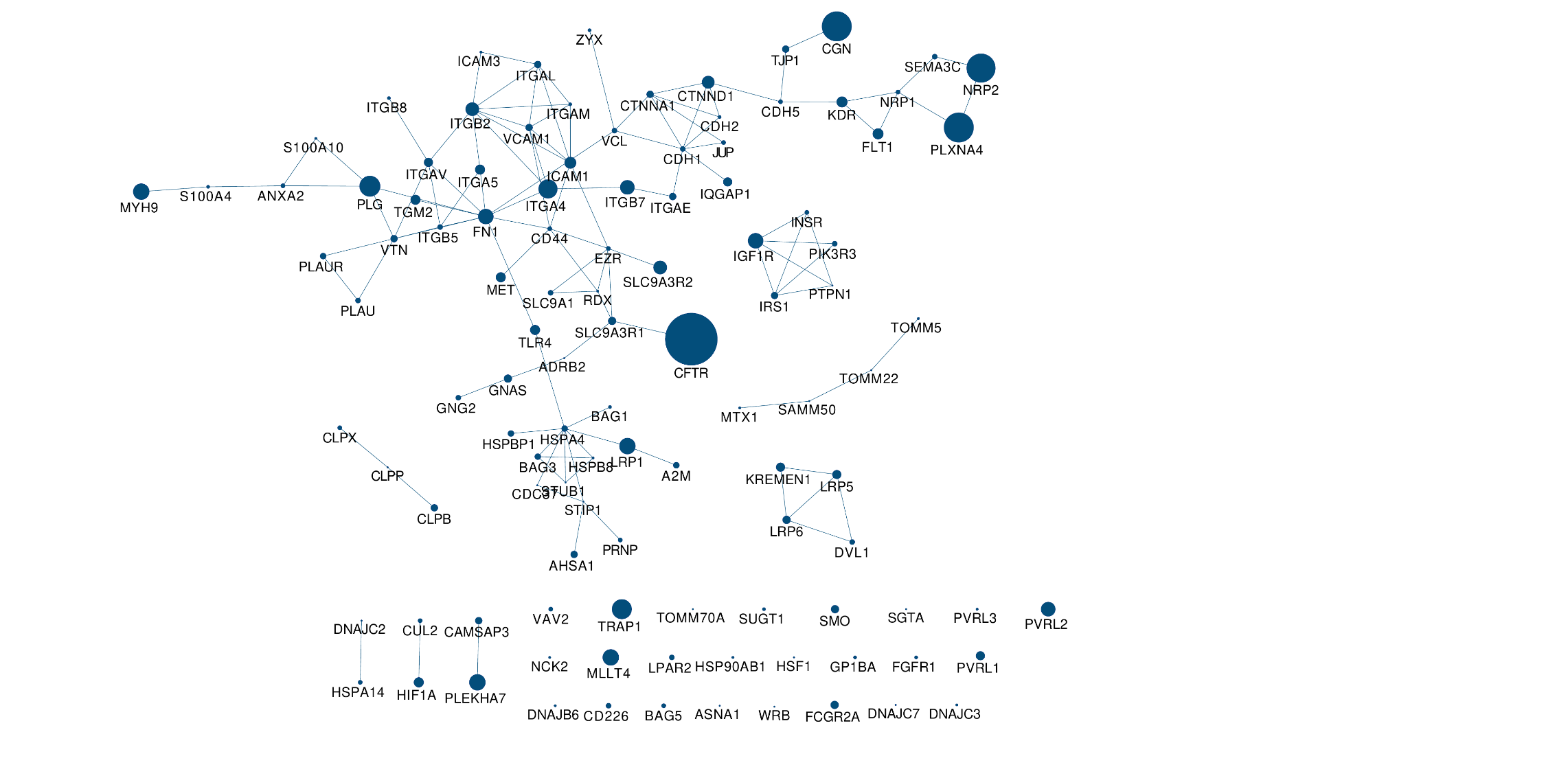

#### **Supplementary Figure 4.** Network representation of Cluster 186, the most significant among the significant clusters with the *Integrin cell surface interactions* as the top-most significant associated pathway. Each node represents a gene in the cluster and the node size indicates the number of ARDS cases harboring qualifying variants in that gene. The lines connecting the nodes represent robust protein-protein interactions according to NHC edge-weighted background network.

#### **Supplementary Figure 5.** Network representation of Cluster 170, the most significant among the significant clusters with the *Toll-like receptor cascades* as the top-most significant associated pathway. Each node represents a gene in the cluster and the node size indicates the number of ARDS patients harboring qualifying variants in that gene. The lines connecting the nodes represent robust protein-protein interactions according to NHC edge-weighted background network.

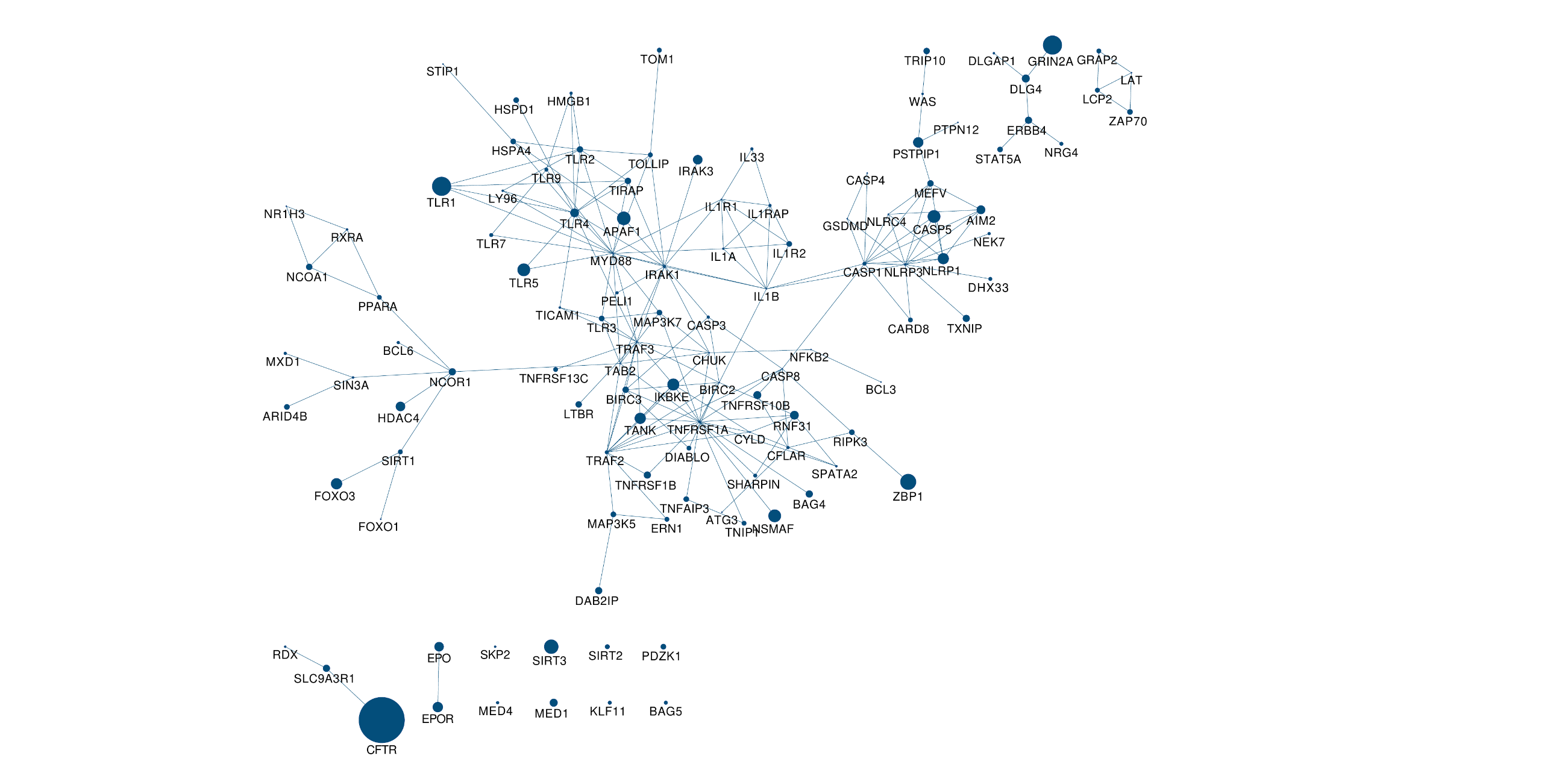

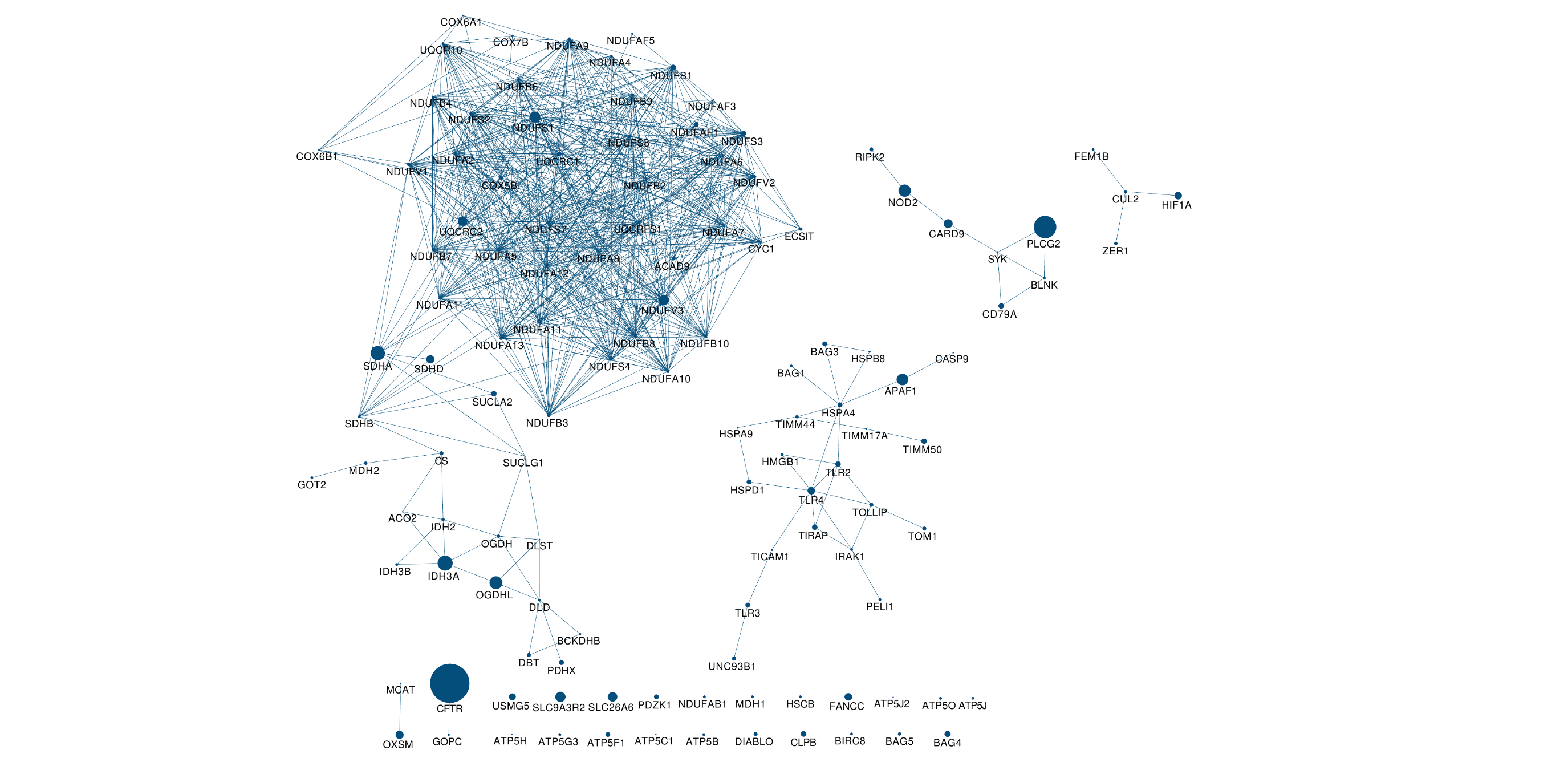

#### **Supplementary Figure 6.** Network representation of Cluster 181, the most significant among the significant clusters with the *Krebs cycle and respiratory electron transport* as the top-most significant associated pathway. Each node represents a gene in the cluster and the node size indicates the number of ARDS patients harboring qualifying variants in that gene. The lines connecting the nodes represent robust protein-protein interactions according to NHC edge-weighted background network.

#### **Supplementary Figure 7.** Network representation of Cluster 169, the only significant cluster with *Transcriptional regulation of white adipocyte differentiation* as the top-most significant associated pathway. Each node represents a gene in the cluster and the node size indicates the number of ARDS patients harboring qualifying variants in that gene. The lines connecting the nodes represent robust protein-protein interactions according to NHC edge-weighted background network.

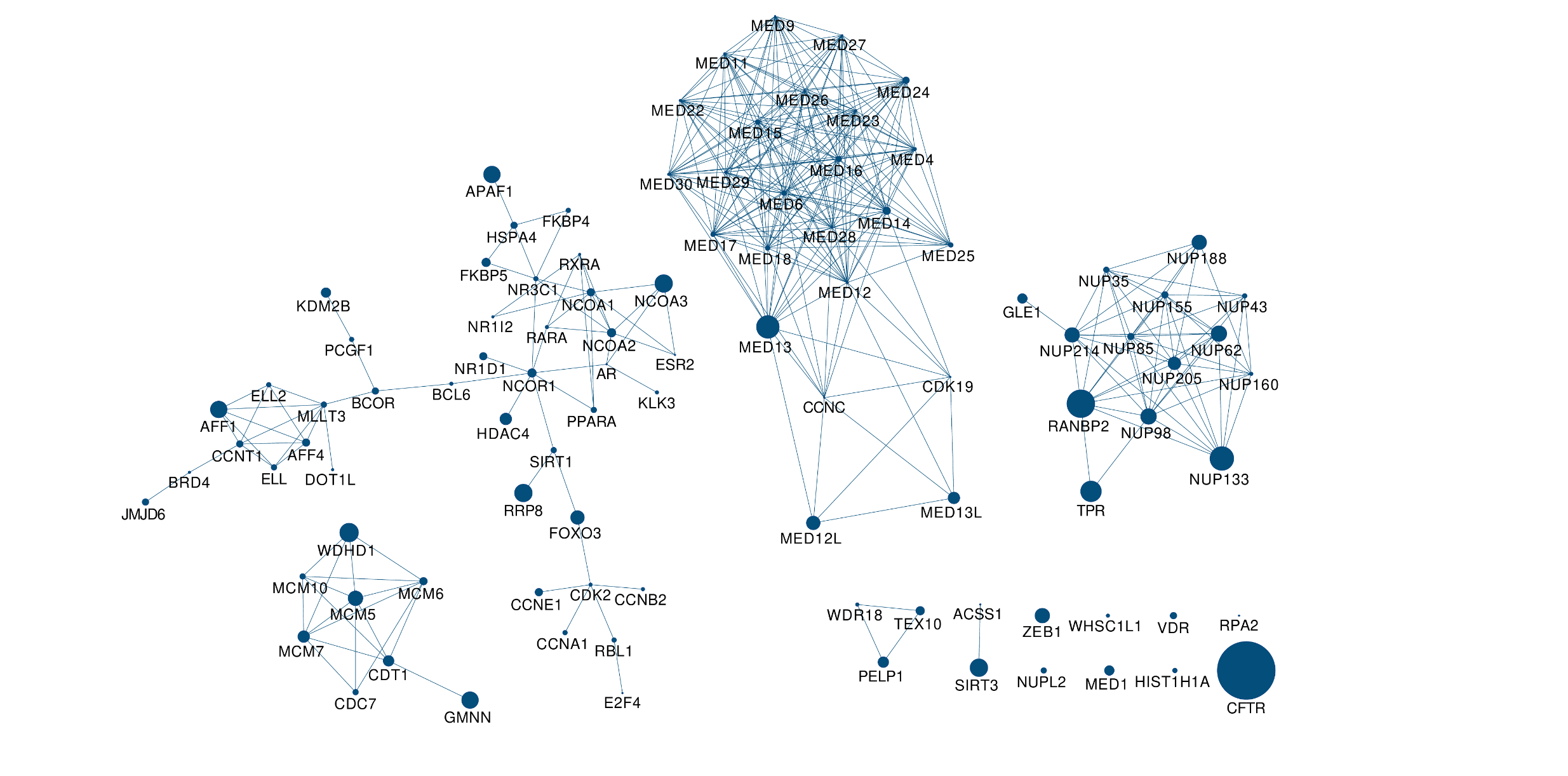

#### **Supplementary Figure 8.** Network representation of Cluster 153, the most significant among the significant clusters with the *Interferon signaling* as the top-most significant associated biological pathway. Each node represents a gene in the cluster and the node size indicates the number of ARDS patients harboring qualifying variants in that gene. The lines connecting the nodes represent robust protein-protein interactions according to NHC edge-weighted background network.

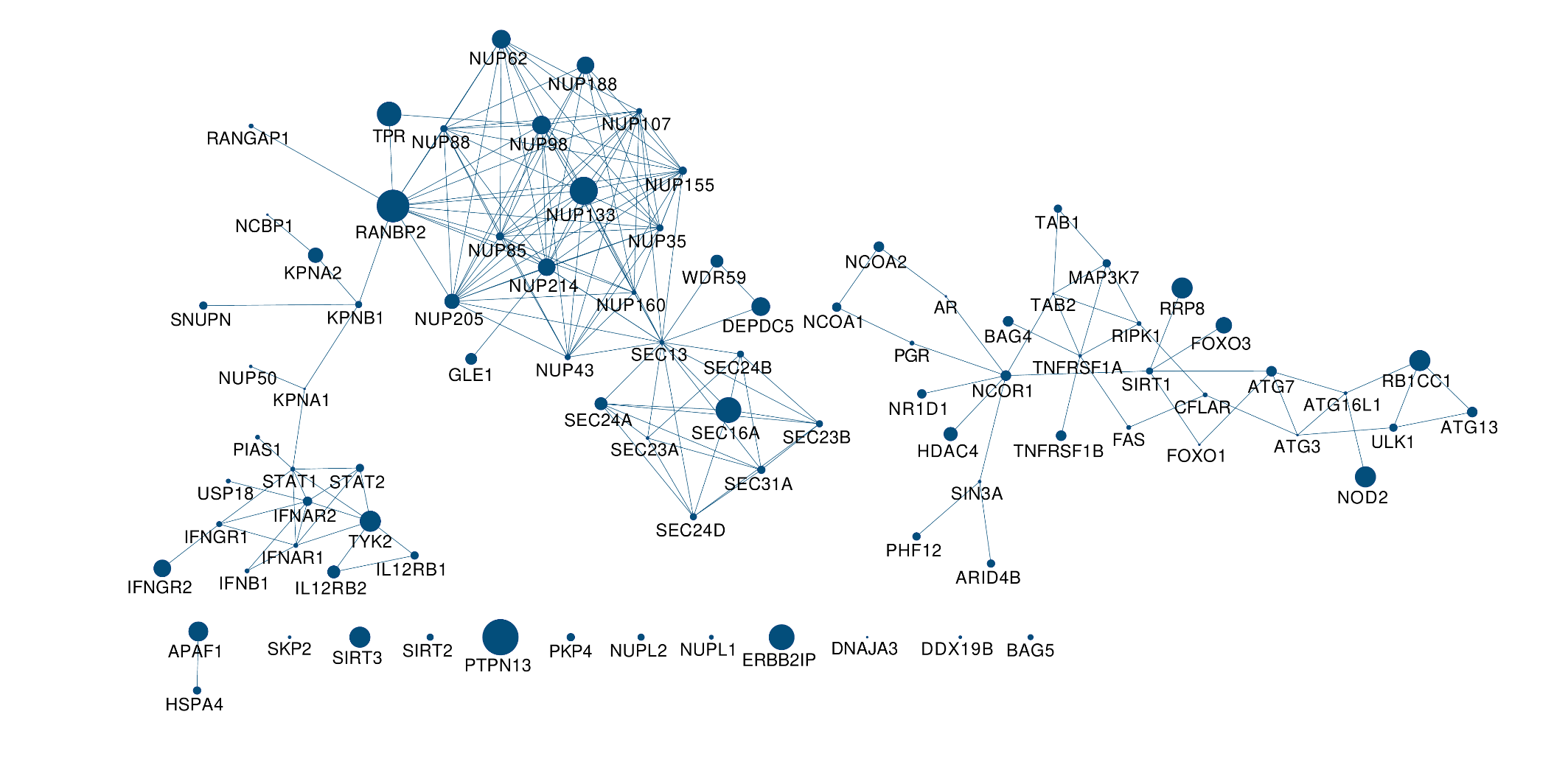

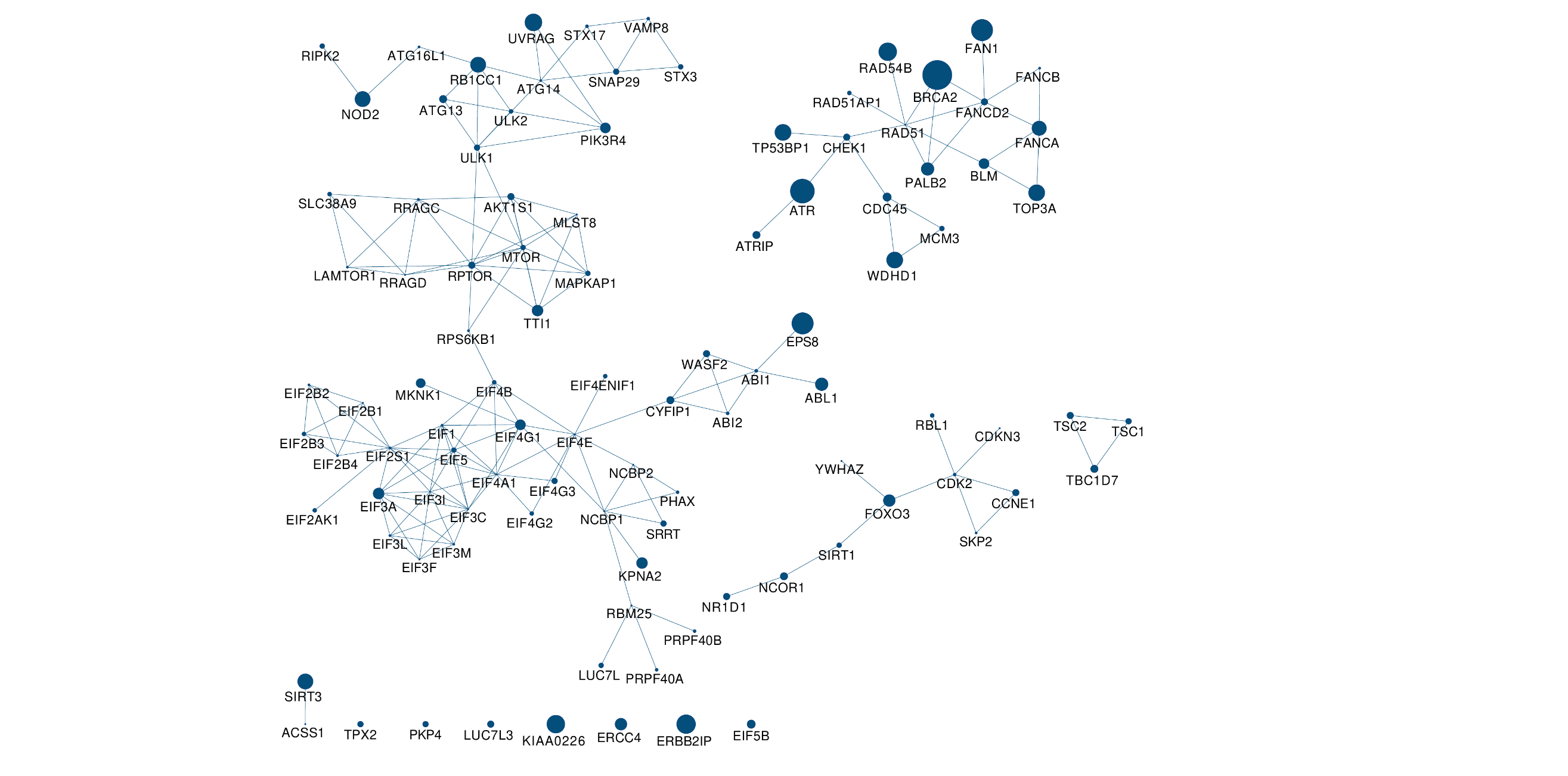

#### **Supplementary Figure 9.** Network representation of Cluster 182, the only significant cluster with *mTOR signaling* as the top-most significant associated pathway. Each node represents a gene in the cluster and the node size indicates the number of ARDS patients harboring qualifying variants in that gene. The lines connecting the nodes represent robust protein-protein interactions according to NHC edge-weighted background network.

#### **Supplementary Figure 10.** Network representation of Cluster 12, the only significant cluster with *Amino acids regulate mTORC1* as the top-most significant associated pathway. Each node represents a gene in the cluster and the node size indicates the number of ARDS patients harboring qualifying variants in that gene. The lines connecting the nodes represent robust protein-protein interactions according to NHC edge-weighted background network.

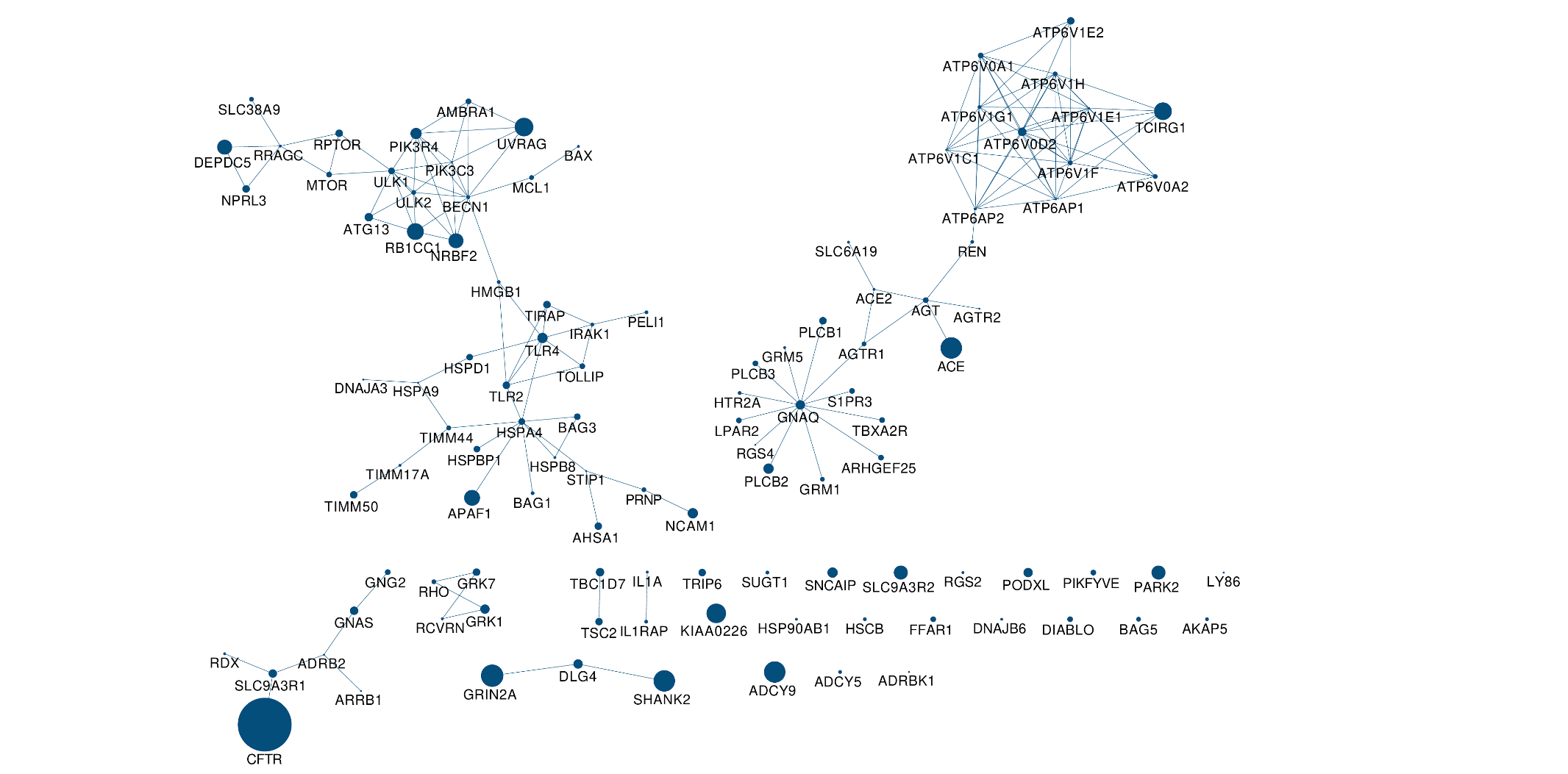

#### **Supplementary Figure 11.** Network representation of Cluster 19, the only significant cluster with *Protein-protein interactions at synapses* as the top-most significant associated pathway. Each node represents a gene in the cluster and the node size indicates the number of ARDS patients harboring qualifying variants in that gene. The lines connecting the nodes represent robust protein-protein interactions according to NHC edge-weighted background network.

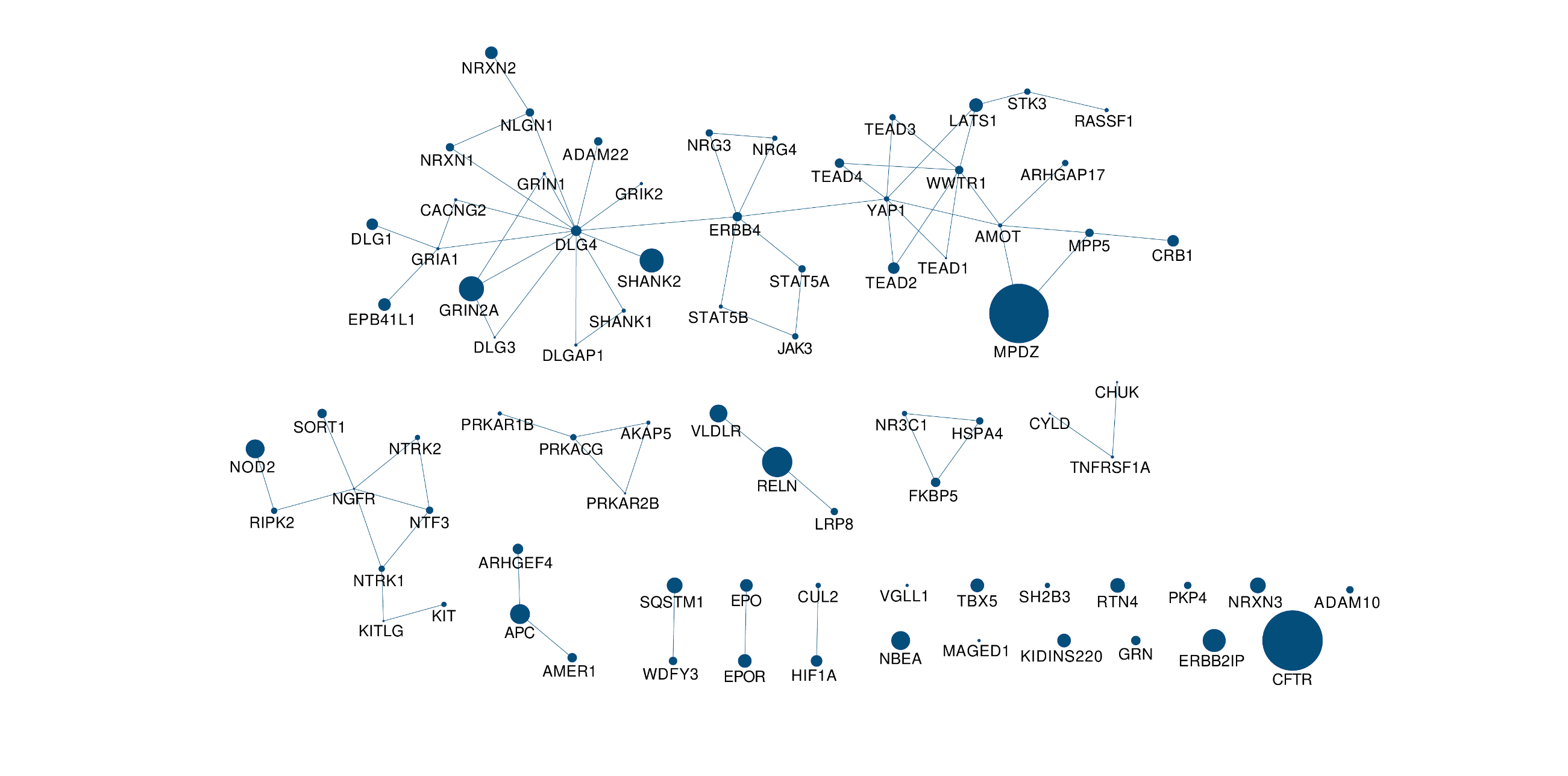

#### **Supplementary Figure 12.** Network representation of Cluster 46, the most significant among the significant clusters with the *mRNA splicing* as the top significant associated pathway. Each node represents a gene in the cluster and the node size indicates the number of ARDS patients harboring qualifying variants in that gene. The lines connecting the nodes represent robust protein-protein interactions according to NHC edge-weighted background network.

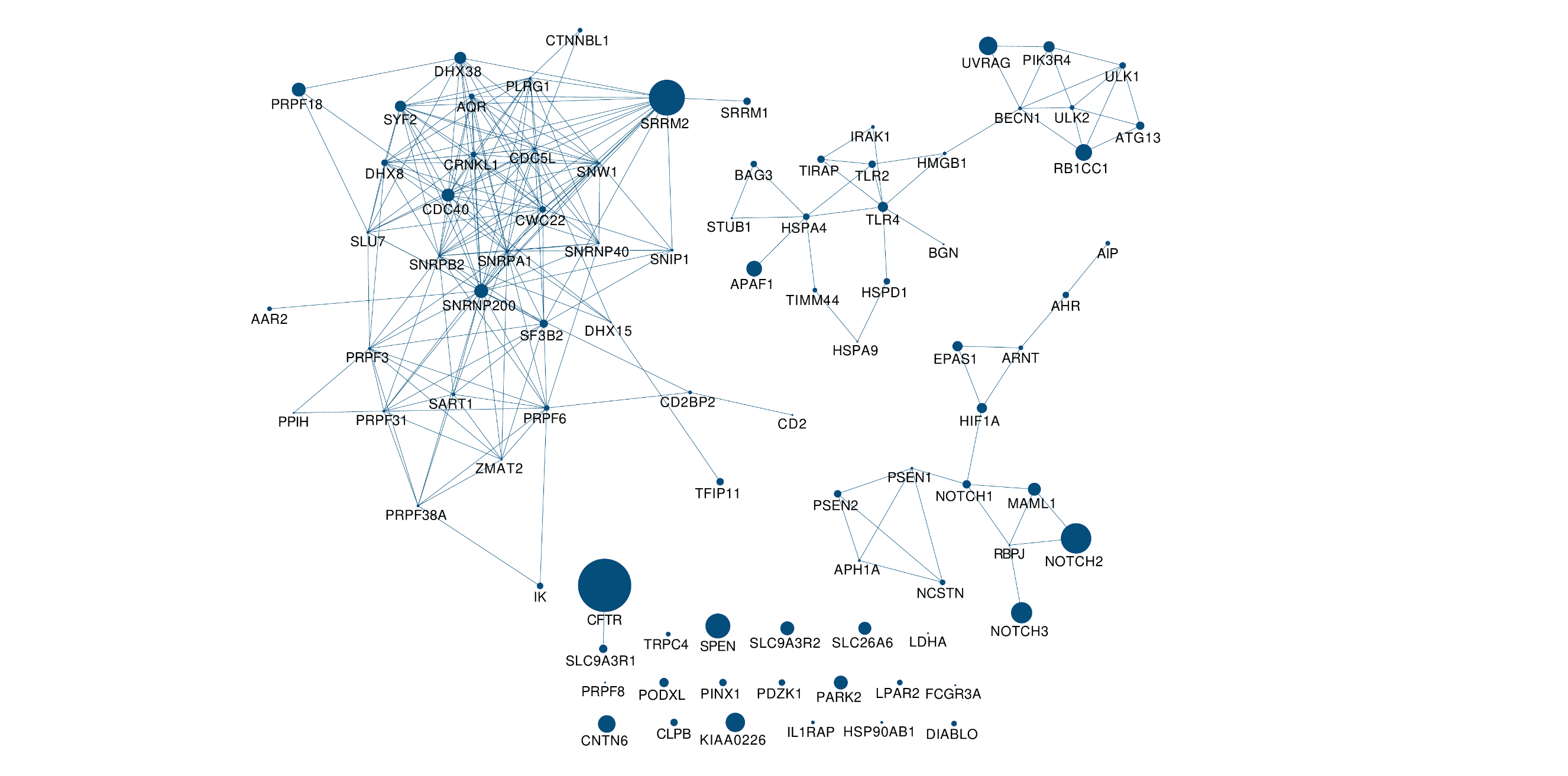

#### **Supplementary Figure 13.** Network representation of Cluster 97, the most significant among the significant clusters with the *DNA repair* as the top-most significant associated pathway. Each node represents a gene in the cluster and the node size indicates the number of ARDS patients harboring qualifying variants in that gene. The lines connecting the nodes represent robust protein-protein interactions according to NHC edge-weighted background network.

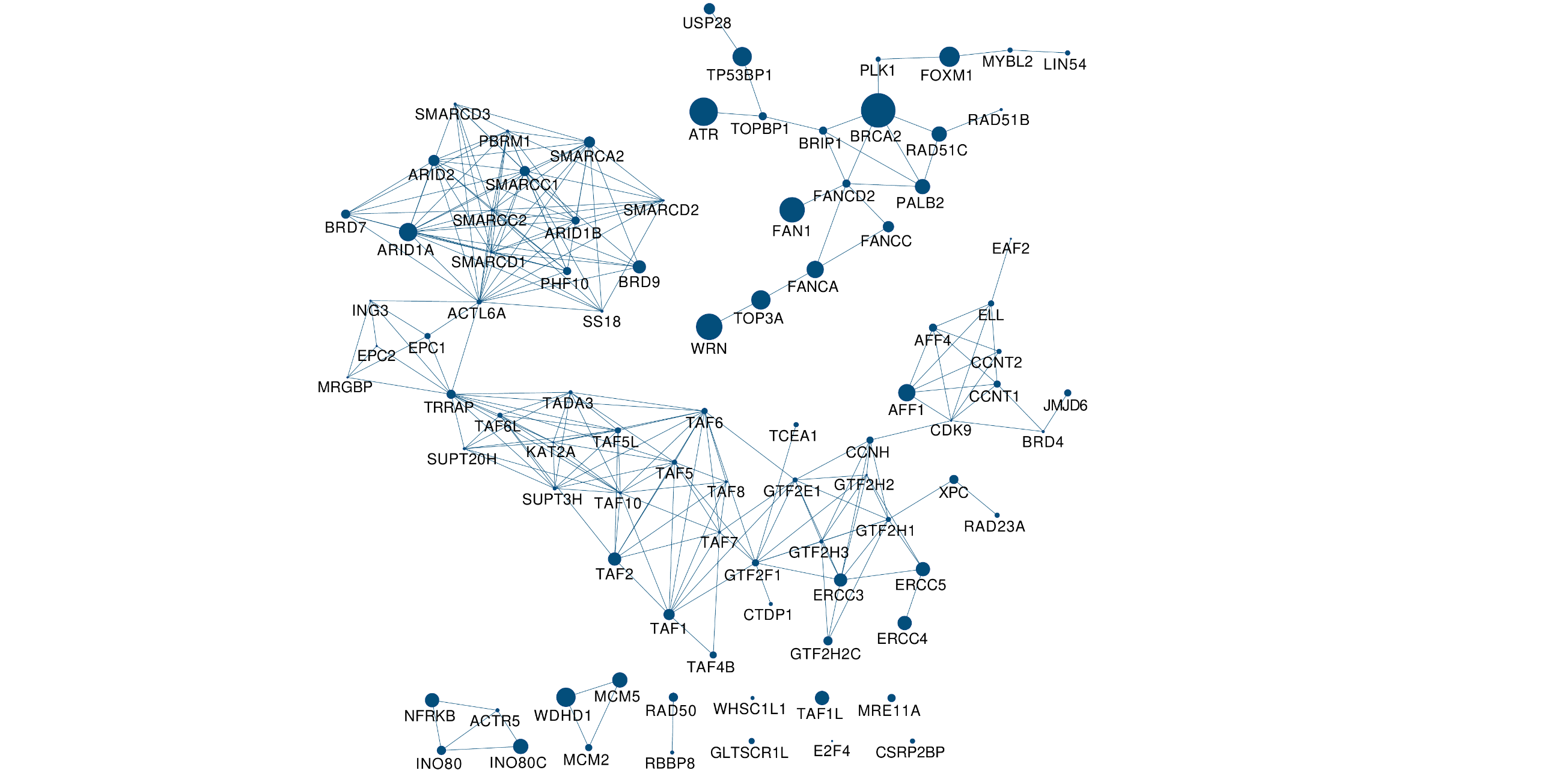
